# From Genomics Data to Causality: An Integrated Pipeline for Mendelian Randomization

**DOI:** 10.1101/2023.11.04.23298053

**Authors:** Jyoti Sharma, Vaishnavi Jangale, Asish Kumar Swain, Pankaj Yadav

**Affiliations:** Department of Bioscience and Bioengineering, Indian Institute of Technology, Jodhpur, 342030, Rajasthan, India; School of Artificial Intelligence and Data Science, Indian Institute of Technology, Jodhpur, 342030, Rajasthan, India

**Keywords:** Mendelian Randomisation, Horizontal Pleiotropy, t-statistics, Heterogeneity, MR-Egger

## Abstract

**Background:** Mendelian randomization (MR) has emerged as a valuable tool for causal inference in genetic epidemiology. Existing MR methods have issues related to pleiotropy and offer limited comprehensiveness. Here, we introduce an integrated MR analysis pipeline designed for GWAS summary statistics data. Our pipeline integrates feature selection, harmonization, and checkpoint mechanisms to improve the accuracy and reliability of MR analysis.

**Methods:** In classical GWAS, the p-value threshold usually does not guarantee to identify causal single-nucleotide polymorphisms (SNPs). In such cases, t-statistics can be considered as imperative and robust criteria for identifying causal SNPs. Therefore, in this study, we computed the t-statistic for all independent SNPs remained after linkage disequilibrium pruning. Next, prior to harmonization, we removed SNPs having a t-statistic below the average t-statistic value. Furthermore, our pipeline incorporates sensitivity analysis tests at each step to reduce the chances of directional pleiotropy.

**Result and Conclusion:** We applied our pipeline to single-sample and two-sample MR study designs, encompassing diverse populations and a wide range of diseases. Our results demonstrate superior performance compared to existing MR methods. In conclusion, our research presents an integrated MR analysis pipeline that significantly enhances the accuracy and reliability of MR studies. By outperforming existing methods and providing comprehensive validation, this pipeline represents a valuable tool for researchers in genetics and epidemiology.

## 1. Introduction

After the completion of gap-less sequencing of the human genome [23], the discovery of genetic variation has become a major area of future research. Comparative genetic studies across ethnically diverse human populations are crucial in understanding the genetic basis of different traits and diseases. These studies aim to understand why certain individuals have a higher susceptibility to specific diseases. Single-nucleotide polymorphisms (SNPs) are of particular importance in such studies due to their high frequency and uniform distribution throughout the genome. Additionally, the density of SNPs needed for mapping complex diseases will likely vary across populations with specific geographical locations. To understand the impact of genetic variants on diseases, it is essential to differentiate between association and causation. Many genome-wide association studies (GWAS) and their statistic models are designed to identify associations between genetic variants and traits or diseases of interest. These associations sometimes could reflect the causal relationship between genetic variants and diseases, but the direction of causality is still not clear. Understanding causal inferences from these GWAS studies can be challenging due to reverse causation, confounding factors, and several other biases[11]. Such issues have been addressed through epidemiological studies by establishing correlations between exposures (e.g., traits, environmental conditions, medication etc.) and outcomes (complex diseases). These studies adopt randomized controlled trials (RCTs), which involve the random assignment of various treatments to individuals of a population. Basically, one “active group” (outcome/disease) that was treated is compared against a “control group” to establish causal relationships. Observational epidemiological studies are not easy to observe results while mediating exposure in RCTs. These studies often present spurious causal relationships between modifiable exposures and disease. In such studies, exposure modification in confined environmental settings underestimates the true association between causal genetic variants and disease. Aside from this, RCTs have a long duration and encounter ethical clearance issues. Moreover, the persistence of confounding and selection bias remains a concern even after the initiation of an RCT. This includes the absence of follow-up data depending on treatment outcomes, potentially resulting in non-randomly missing data issues. However, nowadays, genetically predicted causal inference between exposures and diseases has been popularized in epidemiological studies, which provide new insights into early screening and prevention of disease. However, nowadays, genetically predicted causal inference between exposures and diseases has been popularized in epidemiological studies, which provide new insights into early screening and prevention of disease [20]. The incorporation of genetic variants in epidemiology studies is motivated by the inherent randomization of genetic polymorphism. This means that the genotype of an individual cannot be changed, and interpersonal covariates are normally distributed among the studied population. The key consequence of this randomization is the independent distribution of genetic variations from traits they do not affect. This approach aligns with the principles of Mendel’s laws of segregation and independent assortment [7]. These types of studies are known as Mendelian randomization (MR) studies.

MR studies are an analytical approach that uses such genetic variants as instrumental variables that are robustly associated with the exposure of interest and outcome, whether the effects of the variants on the exposure result in proportional effects on the outcome. MR studies follow three core assumptions which are as follows: i) genetic variants should be associated with exposure, ii) genetic variants should not be associated with confounding factors, and iii) genetic variants should impact outcomes only through exposure. When all of these assumptions are satisfied, MR analysis becomes less susceptible to bias from factors such as non-genetic confounding and reverse causation compared to conventional observational epidemiological analyses [10, 9]. This approach traditionally uses summary statistics datasets of genotype-phenotype associations to evaluate the causal effects of exposures on disease incidence. However, there are research gaps in MR studies, like the potential of methodological because of reverse causation and confounding, which can introduce bias into the results. This limitation impacts the power of an MR analysis in the precision of estimating the genetic association with the outcome, as this association is typically much weaker than the genetic association with the exposures. One of the possible solutions to handle this issue is to use published data on genetic associations with the outcome combined with individual-level data from a cross-sectional study on genetic variants and the exposure to obtain precise MR estimates [6]. The genotyping data from human samples, together with corresponding trait information, involves ethical clearance challenges and is not readily accessible.

Here, we have proposed a pipeline to systematically address the issues of selecting datasets, finding weak causality, and enhancing the efficiency of the genetic instruments. This pipeline incorporates mainly the following steps: the selection and preprocessing of raw data (specifications of datasets), the filtration of genetic instrument variants (feature selection (valid genetic instruments)), the application of MR analysis on selected variants (statistical tests for MR analysis), and the validation of resultant genetic variants (sensitivity analysis). The guidelines from dataset selection to MR analysis should follow all three MR assumptions and check multi-directional pleiotropy to avoid biases that can lead to inaccurate causal estimates. Our proposed pipeline effectively addresses these biases and successfully exhibits a robust framework to analyse causality between exposure and outcome. In classical causality analysis, p-value thresholding usually does not guarantee the identification of causal SNPs. Other than p-values, the selection of valid and effective genetic instruments highly depends on the estimates of exposure and outcome, i.e., *β*_*exposure*_ and *β*_*outcome*_, respectively and their corresponding standard errors (*SE*_*exposure*_ and *SE*_*outcome*_). We applied the filtration method on both exposure and outcome summary statistics datasets using both *β* and *SE*. Moreover, only those genetic instruments exceeding the defined threshold value for these parameters are selected. These selected genetic instruments are then analysed for causality tests by applying different well-established MR methods. The stability of causal estimation in the MR results is evaluated using various sensitivity analysis approaches. To validate the performance of the pipeline, we tested our pipeline on multiple datasets belonging to different exposures and outcomes. Also, to check the robustness of the pipeline, we selected summary statistics datasets from different populations. Summary statistics datasets can be obtained from two types of MR study designs, two-sample MR study and single-sample MR study. The two-sample MR study describes the collection of exposure and outcome datasets from two different sample populations, and the single-sample MR study describes the collection of both exposure and outcome from the same population. For two-sample MR data, we have to collect both datasets from the same super-population. Figure 1 explains the assumptions and study designs of MR analysis. The detailed evaluation of the proposed pipeline is described in the following sections.

**Figure 1:**
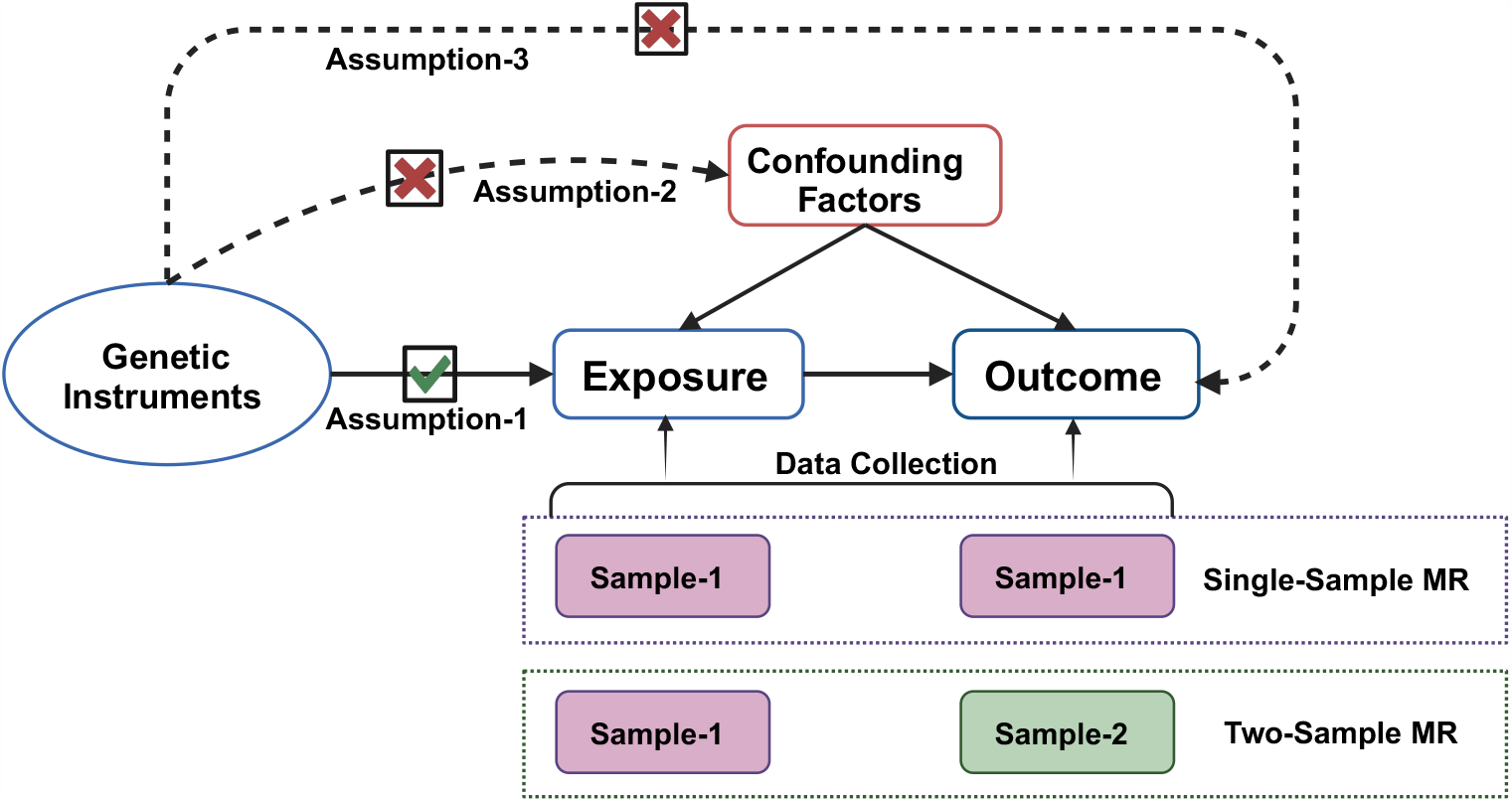
MR assumptions and data collection study designs

## 2. Material and Method

This section provides a comprehensive description of the dataset selection and pre-processing method for summary statistics datasets (step 1), along with the genetic instrument selection (step 2). In addition, various MR approaches are explained with their relative advantages and limitations (step 3), followed by a sensitivity analysis (step 4). The proposed pipeline is shown in Figure 2.

**Figure 2:**
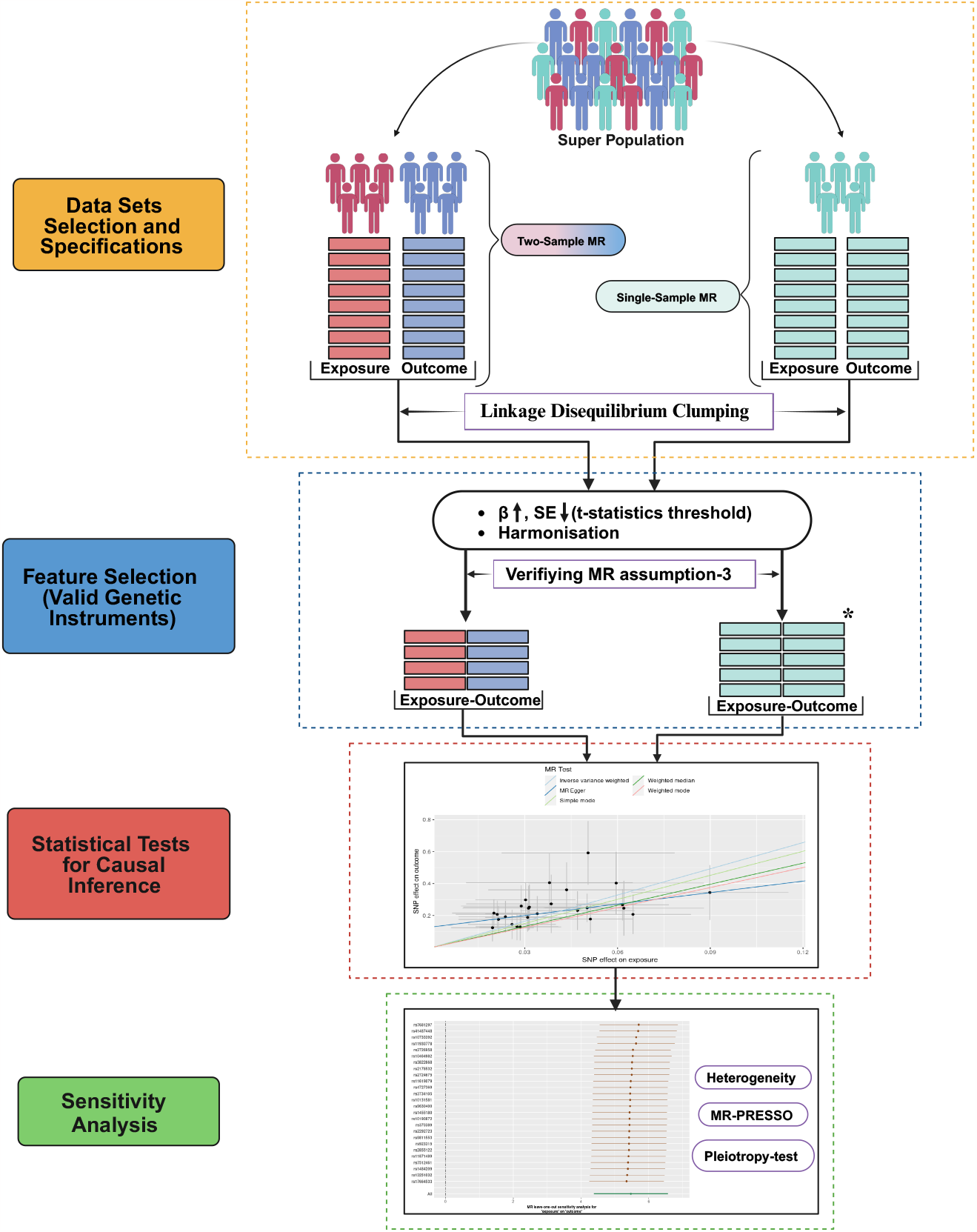
Proposed workflow

### 2.1. Data Sets Selection and Specifications

In this study, the proposed pipeline has been applied to five GWAS summary statistics datasets. Of these, 4 datasets are used for a two-sample MR study design, and one is used for a single-sample MR study design. Before selecting exposure and outcome datasets, it is essential to identify the heritability of the chosen phenotype or trait. Heritability refers to the extent to which the genetic composition of a given sample could be responsible for the variance observed in the phenotype.

#### 2.1.1. Datasets

An overview of all these five datasets is given in Table 1, and the detailed description of all these datasets is as follows:

**Table 1.**
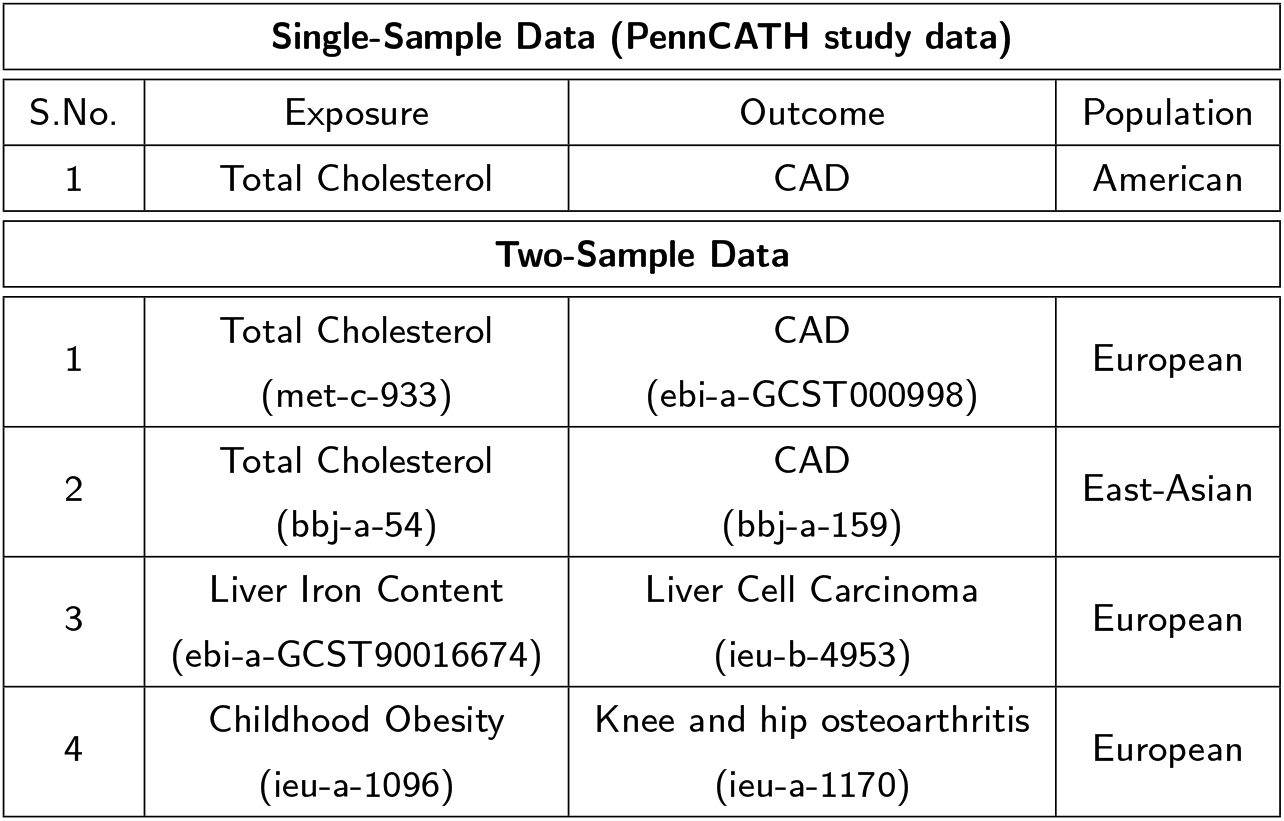
Overview of selected datasets used in this study.

- **Single-sample summary statistic dataset** The genotype and phenotype data for single-sample MR were obtained from the PennCATH cohort study conducted by the University of Pennsylvania’s Medical Center [14]. This dataset comprises a rich array of clinical parameters, including gender, age, CAD status, LDL cholesterol levels, HDL cholesterol levels, and triglycerides. It is anchored by an angiographic CAD case-control GWAS involving 1,401 individuals and 861,473 SNPs. This study removed SNPs with genotype call rates below 100% to keep only high-quality SNPs. Also, SNP with minor allele frequency (MAF <5%) are excluded from genotyping arrays to focus on common SNPs. Population genotype frequency is examined using the inbreeding coefficient and frequency analysis to ascertain the Hardy-Weinberg Equilibrium. Then, high linkage disequilibrium (LD) regions were trimmed to include genetic haplotype block markers. LD was assessed using a *r*^2^ threshold at 0.3 and a 50,000 SNP sliding window. All related samples were eliminated in order to reduce sample biases. After applying all of these pre-processing biological filters, the analysis uses a total of 1277 samples and 41,802 SNPs. The below-discussed two-sample dataset describes the heritability of both exposure and outcome.
- **Two-sample summary statistic datasets**

This study used openly accessible data obtained from the MRC Integrative Epidemiology Unit (IEU, https://gwas.mrcieu.ac.uk/). The datasets are obtained using the R package TwoSampleMR [15]. The following section provides a description of the selected datasets, organized in an “Exposure-Outcome” format.

1. Total Cholesterol-Coronary Heart Disease: The summary-statistics dataset of serum total cholesterol(TC) was obtained from a genome-wide association metaanalysis (GWAS ID: met-c-933). In this dataset, 21,491 samples were analysed, and 118,55,845 SNPs were identified [18]. The dataset of Coronary Heart Disease (CAD) was also obtained from the EBI GWAS Catalog (GWAS ID:ebi-a-GCST000998). In this dataset, 86,995 samples (22,233 cases and 64,762 controls) were analysed with 2,415,020 SNPs [25]. Both exposure and outcome datasets are collected for the European super-population. TC has a heritability of 35-60% [27] and heritability of CAD is approx 40-70% [22].
2. Total Cholesterol-Coronary Heart Disease: The summary-statistics dataset of TC was obtained from the biobank Japan (GWAS ID: bbj-a-54). In this dataset, 128,305 samples were analysed, and 6,108,953 SNPs were identified [16]. The dataset of CAD was also obtained from the biobank Japan (GWAS ID: bbj-a-159). In this dataset, 212,453 samples (29,319 cases and 183,134 controls) were analysed with 24,15,020 SNPs. Both exposure and outcome datasets are collected for the East Asia super-population. The heritability of both exposure and outcome is described in the above dataset detail.
3. Liver Iron Content-Liver Cell Carcinoma: The summary-statistics dataset of liver iron content (LIC) was obtained from EBI GWAS Catalog (GWAS ID: ebi-a-GCST90016674). In this dataset, 32,858 samples were analysed, and 9,275,407 SNPs were identified [19]. The dataset of liver cell carcinoma (LCC) was also obtained from the UK Biobank (GWAS ID: ieu-b-4953). In this dataset, 372,184 samples (168 cases and 372,016 controls) were analysed with 6,304,034 SNPs. Both exposure and outcome datasets are collected for the European super-population. Liver Iron has a heritability of 7% [29], and individual with a family history of Liver Cell Carcinoma has an increased risk of developing this disease [28].
4. Childhood obesity (CO)-Knee and hip osteoarthritis(OA): The summary-statistics data on childhood obesity (CO) were obtained from a genome-wide association meta-analysis (GWAS ID: ieu-a-1096) conducted by the Early Growth Genetics (EGG) consortium [4]. In this dataset, 13,848 children were analysed, and 2,442,739 SNPs were identified. The dataset of Knee and hip osteoarthritis (OA) was also obtained from a genome-wide association meta-analysis (GWAS ID: ieu-a-1170) conducted by the arcOGEN consortium. In this dataset, 14,507 samples (3,498 cases and controls) were analysed with 12,79,483 SNPs. Both exposure and outcome datasets are collected for the European super-population. The estimated heritability of obesity is between 40% and 70% [21]. The estimate of heritability has been reported to be 40% for the knee, 60% for the hip, 65% for the hand, and about 70% for the spine [30].

#### 2.1.2. Datasets pre-processing

The GWAS summary statistics for the single-sample dataset were calculated from linear and logistic regression models for total cholesterol (exposure) and CAD (outcome), respectively. The two-sample MR data are collected with a less-stringent *p*_*value* ≤ 5E-03 to retrieve the maximum possible SNPs. Once the summary statistics data for both exposure and outcome are prepared, the SNPs in linkage disequilibrium (LD) were clumped according to the respective super-population in the 1000 genomes reference panel using a threshold of *r*^2^ ≥ 0 01 and a window size of 10,000 kb with MAF > 0.01.

### 2.2. Feature Selection (Valid Genetic Instruments)

At this step, the first checkpoint is performed to avoid the third assumption of MR, i.e., genetic variants should impact outcomes only through exposure. The clumped SNPs from both datasets, exhibiting a significant correlation with both exposure and outcomes, were excluded to remove the potential confounding effect of horizontal pleiotropy.

The strength of the association of any SNP with the phenotype can be measured through the p-value. However, p-value thresholding overlooked significant SNPs. Therefore, here we consider *β* and the standard error (SE) of corresponding SNPs to understand the impact of SNP on outcome. We calculated the t-statistics as a selection method of valid genetic instruments for both datasets using the following equation:

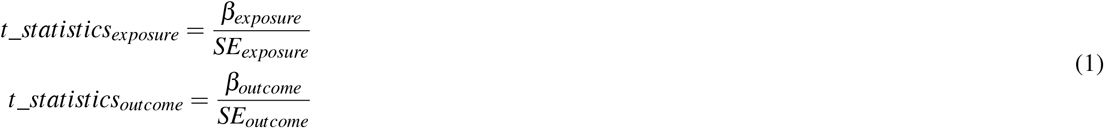

Only those SNPs with t-statistic values greater than the average t-statistics of their respective datasets were selected for subsequent analysis. In addition, the F-statistic was calculated to quantify the strength of selected SNPs as valid genetic instruments for the outcome using the following equation:

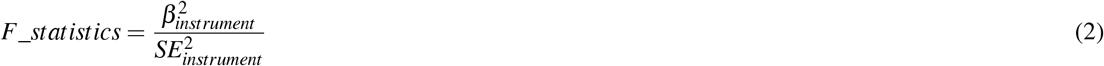

After selecting valid genetic instruments, the resultant SNPs are subsequently harmonised. In the harmonisation process, we attempted to infer all SNPs in the forward-strand.

### 2.3. Statistical Tests for Causal Inference

Statistical tests for MR analysis were performed on R software using TwoSampleMR package. These MR methods included weighted median [2], simple and weighted mode [13], inverse-variance weighted (IVW) [5], and MR-Egger regression [1]. In addition, bias adjustment for MR-Egger with a simulation extrapolation (SIMEX) [3] was also performed.

The weighted median MR method requires that a minimum of 50% of the weight in the analysis derives from valid genetic instruments. The weighted median estimate is insensitive to a pleiotropic genetic variant because the median is unaffected by outliers. In contrast, the mode-based methods (simple mode and weighted mode) require the largest subset of variants that identify the same causal effect for genetic instruments that are valid. Both median and mode-based methods measure the central tendency of variant-specific beta values (causal estimates). These methods are robust to variants with outlier ratio estimates and less impacted by a few pleiotropic variants. For MR analysis, we integrated each SNP effect using IVW, where the estimate was the slope of a zero-intercept regression of SNP-outcome effects on SNP-exposure effects. Besides these methods, MR-Egger can provide unbiased estimates even if all genetic instruments violate the exclusion restriction assumption. However, the genetic instrument must have negligible measurement error (NOME), and the InSIDE (Instrument Strength Independent of Direct Effect) assumption must be satisfied. If the NOME assumption is violated, the MR analysis is performed using the MR-Egger method with simulation extrapolation (SIMEX) correction. While these six methods are best used when genetic instruments consist of a large number of independent SNPs, since *r*^2^ values between SNPs in our instruments were low (*r*^2^ *>* 0.001), we included them with a further sensitivity analysis to account for correlation in the MR Egger analysis. While these six methods are best used when genetic instruments consist of a large number of independent SNPs, since *r*^2^ values between SNPs in our instruments were low (*r*^2^ *>* 0.001), we included them with a further sensitivity analysis to account for correlation in the MR Egger analysis.

### 2.4. Sensitivity Analysis

Before conducting the sensitivity analysis of the resultant SNPs, the third checkpoint is enacted, which examines reverse causality. A directionality test is carried out to explore the potential existence of reverse causality within the chosen datasets to reduce the risk of spurious causal associations. For this purpose “directionality_test” function is used from the “TwoSampleMR” package.

In MR studies, a heterogeneity test refers to a statistical evaluation that examines the compatibility between instrumental variable estimations using each genetic variant. This evaluation is known as an over-identification test because it identifies the same causal effect through every instrumental variant. The presence of heterogeneity signals potential issues, either indicating violations of modeling assumptions or suggesting that some genetic variants are not conforming to the instrumental variant assumptions. Cochran’s Q is a statistical measure used to quantify heterogeneity. It is computed by summing the squared differences between the individual study effects and the pooled effect across studies, with the weights being consistent with the pooling method.

Heterogeneity indicates the possibility that different causal mechanisms may contribute to the same disorder. Moreover, it may occur when genetic polymorphisms linked to the exposure of interest directly influence the outcome through various pathways, potentially leading to biased estimates of causal effects [12]. Further, we performed the MR-PRESSO (Mendelian Randomization Pleiotropy RESidual Sum and Outlier) test [26]. In this test, genetic variants are systematically removed based on their contributions to heterogeneity, underscoring the significance of addressing heterogeneity in causal inference.

However, it is noteworthy that MR-PRESSO has its limitations. In some cases where outlier removal or covariate adjustment cannot entirely eliminate horizontal pleiotropy, additional pleiotropy tests become imperative to ensure the accurate study of the causal relationship between exposure and outcome. One such test is the “pleiotropy test” from twosampleMR package. It involves MR-Egger regression analysis as its regression intercept can assess the magnitude of pleiotropy. The intercept closer to zero indicates a lower likelihood of genetic pleiotropy [17]. The p-value ≥ 0.05 indicates that there is no significant horizontal pleiotropy in the genetic instruments used in the analysis.

In order to assess the specific impact of each SNP on the causal relationship, we employ a Leave-one-out analysis [8]. Additionally, we utilize forest plots to evaluate the effect estimates of genetic variation.

## 3. Results

The proposed method focuses on selecting valid genetic instruments to improve the precision of causative SNPs using different MR methods. The results for both types of MR study designs are described in this section.

- **Single-sample summary statistic dataset** On the single-sample dataset, the LD clumping step produced 4093 SNPs and 4081 SNPs for the exposure and outcome datasets, respectively. There was no shared SNP that was found to be significantly associated with both exposure and outcome, and it validated the follow-up of the third MR assumption. Next, the average t-statistic threshold was applied, which resulted in a total of 988 SNPs and 959 SNPs in the exposure and outcome dataset, respectively. After this, data harmonization was performed on these filtered SNPs, resulting in a total of 26 SNPs. F statistics were calculated to estimate the sample overlap effect and weak instrument bias considering the relatively relaxed threshold, and an F > 10 was considered dubious bias [24]. However, as the single-sample data has a low sample size, the F statistics of all SNPs follow the standard statistical threshold (i.e. > 3.95). Afterward, all six MR methods were applied to analyze this harmonized dataset. All MR approaches demonstrated a statistically significant causal association between total cholesterol and CAD, except MR Egger. The classic IVW model IVW was employed in the primary MR analyses. When directional pleiotropy is absent, the IVW method can deliver a relatively stable and accurate causal evaluation by using a meta-analytic approach to combine Wald estimates for each IV. Table 2 displayed the detailed results of all MR methods. Figure 3 presents a scatter plot of different MR methods, depicting the positive correlation between TC and the probability of developing CAD.

**Table 2.** MR results of single-sample dataset after t-statistics filter.

| MR methods | #SNP | $\beta$ | SE | p-value |
| --- | --- | --- | --- | --- |
| MR-Egger | 26 | 2.3926 | 1.3928 | 9.87E-02 |
| Weighted median | 26 | 4.3987 | 1.0127 | 1.40E-05 |
| Inverse variance weighted | 26 | **5.4731** | **0.5579** | **1.02E-22** |
| Simple mode | 26 | 5.0182 | 1.4023 | 1.45E-03 |
| Weighted mode | 26 | 4.1684 | 1.0162 | 3.81E-04 |
| MR-Egger SIMEX | 26 | 2.827415 | 1.479024 | 6.792946E-02 |

**Figure 3:**
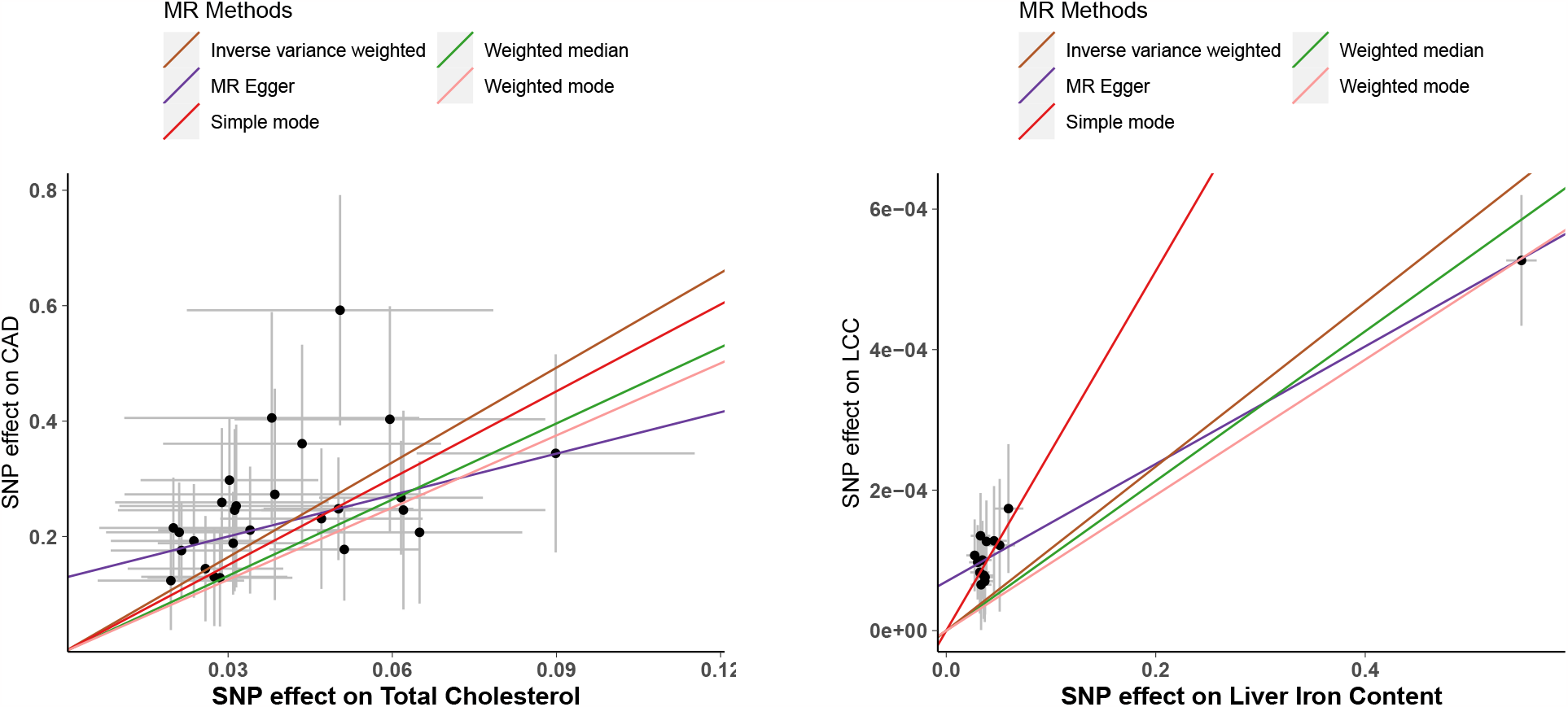
Scatter plot of different MR methods for single-sample and two-sample data

**Figure 4:**
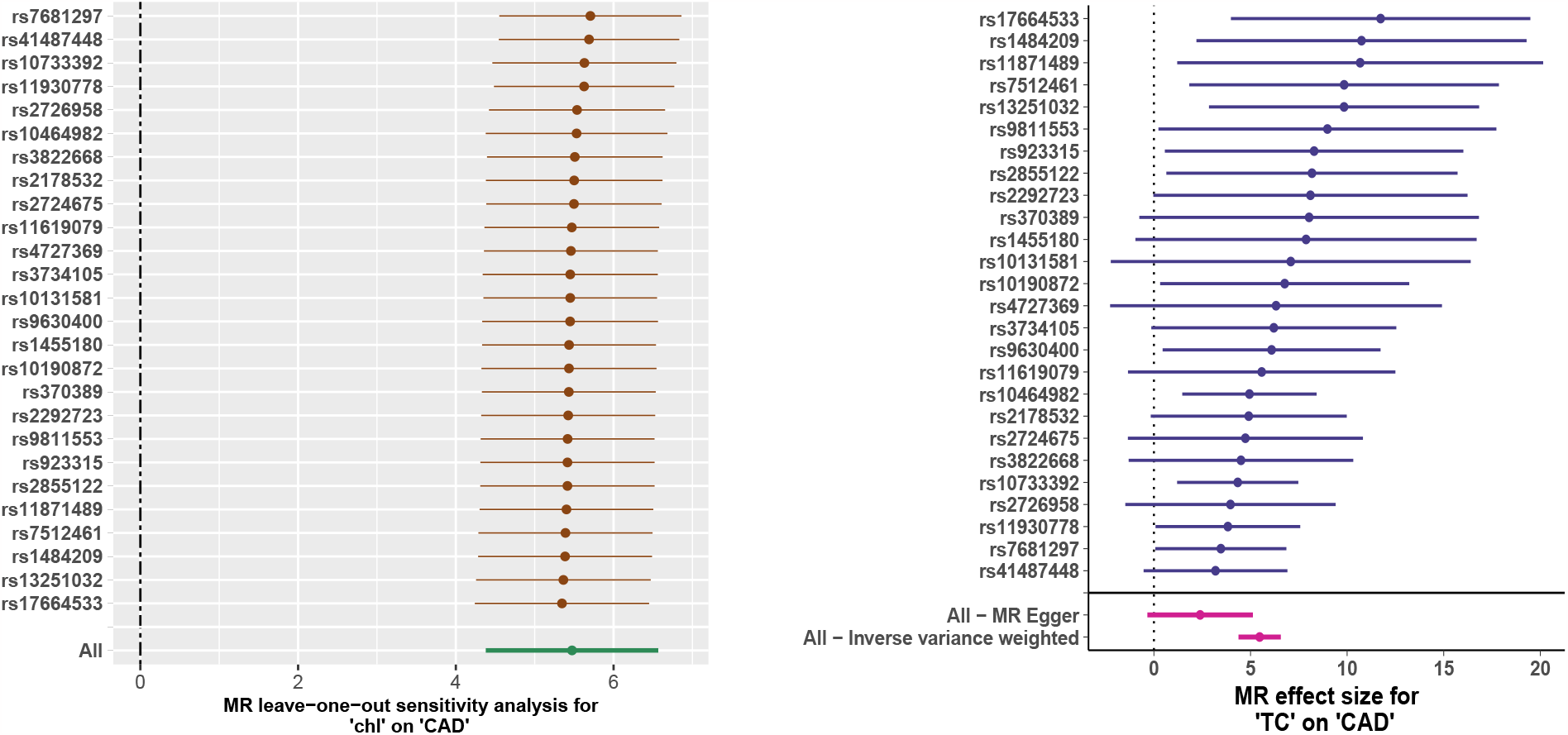
Sensitivity analysis of Single-Sample MR (Leave-one-out and Forest plot)

**Figure 5:**
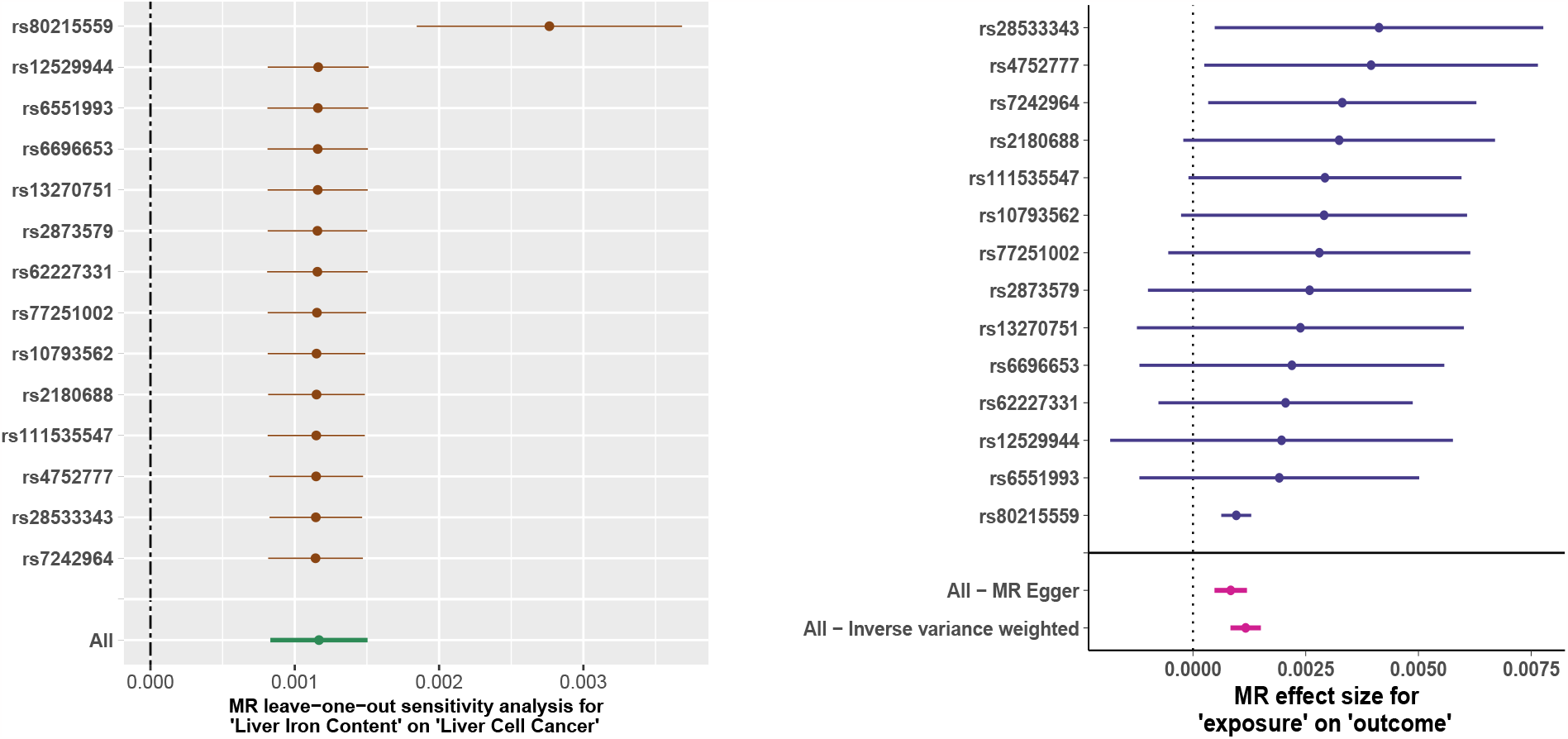
Sensitivity analysis of Two-Sample MR (Leave-one-out and Forest plot) Further, the directionality of a causal relationship is evaluated, and the directionality test displays the direction of causality from the TC, i.e., exposure to CAD, i.e., outcome. This analysis indicates the absence of reverse causality in the single-sample dataset. Following the analysis, the heterogeneity test yielded non-significant findings, as shown by Cochran’s Q value (= 9.6) being considerably lower than the degrees of freedom (= 24) and a p-value of 0.9959. These results suggest the absence of heterogeneity. After the heterogeneity, we checked probable outliers in the selected genetic instrument using MR-PRESSO. Our analysis did not identify any statistically significant outliers, suggesting that the probability of horizontal pleiotropy is very minimal. The pleiotropy test obtained a p-value of 0.0237, which does not reach statistical significance about the existence of horizontal pleiotropy. The intercept value of 0.12 is also minor, suggesting the lack of multiple pleiotropic effects in the analyzed single-sample dataset. If the default parameters are applied and t-statistics is not applied, the single-sample dataset shows insignificant results. After the harmonisation, 679 SNPs are obtained, and out of 679 SNPs, 668 SNPs are involved in MR analysis. The IVW method estimated the coefficient (*β*) −0.0742 with a standard error of 0.2349, and the p-value is 0.7520. The results are not significant and also show a spurious negative association between TC and CAD.
- **Two-sample summary statistic datasets** The two-sample dataset is collected from the database of MRC IEU with p-value filter ≥ 5E-03 and *r*^2^ = 0.01. This section presents a comprehensive analysis of one of our selected datasets, the impact of LIC on LCC. The results of additional datasets are presented in a supplementary document. A total of 4233 SNPs and 1624 SNPs were obtained for the LIC and LCC datasets, respectively. There was no shared SNP that was found to be significantly associated with both exposure and outcome, and it validated the follow-up of the third MR assumption. After pruning these datasets according to their LD values, the average t-statistic threshold was applied, which resulted in a total of 473 SNPs and 221 SNPs in the exposure and outcome datasets, respectively. After this, data harmonization was performed on these filtered SNPs, resulting in a total of 14 SNPs. F statistics were calculated to estimate the sample overlap effect and weak instrument bias considering the relatively relaxed threshold, and an F > 10 was considered dubious bias [24]. Here we have sufficiently large size of samples, and the F statistics of all SNPs are above 10, thus indicating their strong candidacy for causality tests. Afterward, all six MR methods were applied to analyze this harmonized dataset. All MR approaches demonstrated a statistically significant causal association between iron content in the liver and LCC, including MR Egger. Table 3 shows the detailed results of all MR methods. Figure 3 presents a scatter plot of different MR methods, depicting the positive correlation between iron contents in the liver and the probability of developing LCC.

**Table 3.** MR results of two-sample dataset after t-statistics filter(liver cancer) Further, the directionality test is performed, and it displays the direction of causality from the iron content, i.e., exposure to LCC, i.e., outcome. It indicates that no reverse causality is present in this two-sample dataset. Subsequently, heterogeneity is performed, and the value of Cochran’s Q is 1.9898, which is much lower than the degrees of freedom (i.e., 12 for this dataset), and the p-value is not statistically significant (= 0.9994). This result indicates the absence of heterogeneity. After the heterogeneity, we checked potential outliers in the selected genetic instrument using MR-PRESSO, and we didn’t detect any significant outliers here, indicating that the likelihood of horizontal pleiotropy is extremely low. Finally, the pleiotropy test is conducted, and the obtained p-value is 3.49E-03, which is significant for the presence of horizontal pleiotropy; however, the intercept value is extremely small, 6.98E-05, which has no effect on the analysis as a whole. With the default parameters, LIC and LCC are obtained from the MRC IEU database. A total of 10 SNPs and 8 SNPs are retrieved for LIC and LCC, respectively. Following the pruning of SNPs in LD, the harmonisation process was performed. After the harmonisation, a total of 5 SNPs are found, which are involved in MR analysis. The IVW method estimated the *β* 8.42E-04 with a standard error of 2.50E-03 and the p-value 7.58E-04. The results are comparatively less significant with a very small number of genetic instruments.

| Method | #SNP | $\beta$ | SE | p-value |
| --- | --- | --- | --- | --- |
| MR-Egger | 14 | 8.36E-04 | 1.83E-04 | 6.57E-04 |
| Weighted median | 14 | 1.06E-03 | 1.65E-04 | 1.31E-10 |
| Inverse variance weighted | 14 | 1.16E-03 | 1.71E-04 | 1.06E-11 |
| Simple mode | 14 | 2.55E-03 | 1.13E-03 | 4.23E-02 |
| Weighted mode | 14 | 9.64E-04 | 1.74E-04 | 9.61E-05 |
| MR-Egger SIMEX | 14 | 8.33E-04 | 1.43E-04 | 8.41E-05 |

### 3.1. Performance evaluation

To evaluate the effectiveness and robustness of our proposed pipeline, we applied it to different datasets of twosample data collected from varied super-populations. In Table 4, we present a comparison of p-values between two frequently used MR methods, considering both the proposed pipeline and the standard default parameter settings implemented in MR analysis across all the specified datasets.

**Table 4.** Comparative MR results of two-sample datasets before and after t-statistics filter.

| MR study Design | Dataset | With default parameters |  | After Proposed pipeline |  |
| --- | --- | --- | --- | --- | --- |
|  |  | IVW | MR-Egger | IVW | MR-Egger |
| Single-Sample | TC-CAD (American) | 0.752 | 0.6939 | 1.02E-22 | 9.87E-02 |
| Two-Sample | TC-CAD (European) | 1.25E-04 | 0.1184 | 5.04E-06 | 3.32E-03 |
|  | TC-CAD (East-Asian) | 1.84E-11 | 1.59E-05 | 1.31E-31 | 7.08E-07 |
|  | Liver iron-LCC (European) | 7.58E-04 | 5.10E-02 | 1.06E-11 | 6.57E-04 |
|  | CO-OA (European) | 1.9E-03 | 0.3483 | 1.68E-16 | 2.60E-02 |

## 4. Conclusion

This work introduces an optimized MR analysis pipeline that effectively elucidates a causal association between exposure and outcome within various super populations while minimizing susceptibility to horizontal pleiotropy and outlier effects. Although the MR-Egger method has the potential to address directional pleiotropy and yield causal estimates, its statistical power remains somewhat constrained. Horizontal pleiotropy represents a pivotal challenge to MR analysis, as it can distort causal estimations, diminish statistical power, and potentially lead to spurious positive causal connections. However, by implementing bias correction through SIMEX, we were able to unveil a substantial and statistically significant causal relationship between exposure and outcome while mitigating the likelihood of directional pleiotropic influence.

Furthermore, conventional p-value thresholding techniques may not consistently identify SNPs with significant causal associations within GWAS summary statistics datasets. In such scenarios, t-statistics emerge as a robust alternative for selecting SNPs as potential genetic instruments. In single-sample MR datasets, both exposure and outcome variables are derived from the same sample, which eases the data harmonization step. Nevertheless, it is accompanied by the potential drawback of weak genetic instruments. To counteract this limitation, our proposed pipeline incorporates sensitivity analysis tests at each stage and employs t-statistics thresholding to mitigate the influence of weak bias. To validate our approach, we calculated F-statistics. Despite the limited sample size in our study, F-statistics exceeded the threshold of 10 for two-sample datasets.

In summary, the proposed pipeline extends beyond single-sample MR analysis and demonstrates its versatility by successfully handling two-sample data, even in cases where sample dissimilarity is a concern. Our pipeline excelled in both the selection of valid genetic instruments and the detection of robust causal relationships. Thus, our study identified specific SNPs with a causal relationship to outcome. We anticipate that our proposed pipeline will exhibit robust performance when applied to high-dimensional datasets from diverse populations, thus offering a valuable tool for future MR investigations.

## Data availability statement

### Ethical Approval

This research is conducted on summary statistics from published studies and publicly available GWAS data. Patient consent and ethical approval for publication are not applicable.

## Data Availability

All data produced are available online at MRC Integrative Epidemiology Unit database and github.

https://gwas.mrcieu.ac.uk/

## Acknowledgements

This work is partially supported by the GenomeIndia Project, Department of Biotechnology, India and the Ministry of Education, India.

## References

[1] Bowden, J., Davey Smith, G., Burgess, S., 2015. Mendelian randomization with invalid instruments: effect estimation and bias detection through egger regression. International journal of epidemiology 44, 512–525.

[2] Bowden, J., Davey Smith, G., Haycock, P.C., Burgess, S., 2016a. Consistent estimation in mendelian randomization with some invalid instruments using a weighted median estimator. Genetic epidemiology 40, 304–314.

[3] Bowden, J., Del Greco M F., Minelli, C., Davey Smith, G., Sheehan, N.A., Thompson, J.R., 2016b. Assessing the suitability of summary data for two-sample mendelian randomization analyses using mr-egger regression: the role of the i 2 statistic. International journal of epidemiology 45, 1961–1974.

[4] Bradfield, J.P., Vogelezang, S., Felix, J.F., Chesi, A., Helgeland, Ø., Horikoshi, M., Karhunen, V., Lowry, E., Cousminer, D.L., Ahluwalia, T.S., et al., 2019. A trans-ancestral meta-analysis of genome-wide association studies reveals loci associated with childhood obesity. Human molecular genetics 28, 3327–3338.

[5] Burgess, S., Butterworth, A., Thompson, S.G., 2013. Mendelian randomization analysis with multiple genetic variants using summarized data. Genetic epidemiology 37, 658–665.

[6] Burgess, S., Scott, R.A., Timpson, N.J., Davey Smith, G., Thompson, S.G., Consortium, E.I., 2015. Using published data in mendelian randomization: a blueprint for efficient identification of causal risk factors. European journal of epidemiology 30, 543–552.

[7] Burgess, S., Smith, G.D., Davies, N.M., Dudbridge, F., Gill, D., Glymour, M.M., Hartwig, F.P., Kutalik, Z., Holmes, M.V., Minelli, C., Morrison, J.V., Pan, W., Relton, C.L., Theodoratou, E., 2023. Guidelines for performing mendelian randomization investigations: update for summer 2023. Wellcome Open Research 4, 186. URL: https://doi.org/10.12688/wellcomeopenres.15555.3, doi:10.12688/wellcomeopenres.15555.3.

[8] Cheng, H., Garrick, D.J., Fernando, R.L., 2017. Efficient strategies for leave-one-out cross validation for genomic best linear unbiased prediction. Journal of animal science and biotechnology 8, 1–5.

[9] Davey Smith, G., Hemani, G., 2014. Mendelian randomization: genetic anchors for causal inference in epidemiological studies. Human Molecular Genetics 23, R89–R98. URL: 10.1093/hmg/ddu328, doi:10.1093/hmg/ddu328, arXiv:https://academic.oup.com/hmg/article-pdf/23/R1/R89/9459019/ddu328.pdf.

[10] Davies, N.M., Holmes, M.V., Davey Smith G., 2018. Reading mendelian randomisation studies: a guide, glossary, and checklist for clinicians. BMJ 62. URL: https://www.bmj.com/content/362/bmj.k601, doi:10.1136/bmj.k601, arXiv: https://www.bmj.com/content/362/bmj.k601.full.pdf.

[11] Ebrahim, S., Davey Smith G., 2008. Mendelian randomization: can genetic epidemiology help redress the failures of observational epidemiology? Human genetics 123, 15–33.

[12] Greco M F.D., Minelli, C., Sheehan, N.A., Thompson, J.R., 2015. Detecting pleiotropy in mendelian randomisation studies with summary data and a continuous outcome. Statistics in medicine 34, 2926–2940.

[13] Hartwig, F.P., Davey Smith, G., Bowden, J., 2017. Robust inference in summary data mendelian randomization via the zero modal pleiotropy assumption. International journal of epidemiology 46, 1985–1998.

[14] Helgadottir, A.e.a., 2006. A variant of the gene encoding leukotriene a4 hydrolase confers ethnicity-specific risk of myocardial infarction. Nature genetics 38, 68–74.

[15] Hemani, G., Zheng, J., Elsworth, B., Wade, K., Baird, D., Haberland, V., Laurin, C., Burgess, S., Bowden, J., Langdon, R., Tan, V., Yarmolinsky, J., Shibab, H., Timpson, N., Evans, D., Relton, C., Martin, R., Davey Smith, G., Gaunt, T., Haycock, P., The MR-Base Collaboration, 2018. The mr-base platform supports systematic causal inference across the human phenome. eLife 7, e34408. URL: https://elifesciences.org/articles/34408, doi:10.7554/eLife.34408.

[16] Kanai, M., Akiyama, M., Takahashi, A., Matoba, N., Momozawa, Y., Ikeda, M., Iwata, N., Ikegawa, S., Hirata, M., Matsuda, K., et al., 2018. Genetic analysis of quantitative traits in the japanese population links cell types to complex human diseases. Nature genetics 50, 390–400.

[17] Kemp, J.P., Sayers, A., Smith, G.D., Tobias, J.H., Evans, D.M., 2016. Using mendelian randomization to investigate a possible causal relationship between adiposity and increased bone mineral density at different skeletal sites in children. International journal of epidemiology 45, 1560–1572.

[18] Kettunen, J., Demirkan, A., Würtz, P., Draisma, H.H.M., Haller, T., Rawal, R., Vaarhorst, A., Kangas, A.J., Lyytikäinen, L.P., Pirinen, M., Pool, R., Sarin, A.P., Soininen, P., Tukiainen, T., Wang, Q., Tiainen, M., Tynkkynen, T., Amin, N., Zeller, T., Beekman, M., Deelen, J., van Dijk, K.W., Esko, T., Hottenga, J.J., van Leeuwen, E.M., Lehtimäki, T., Mihailov, E., Rose, R.J., de Craen, A.J.M., Gieger, C., Kähönen, M., Perola, M., Blankenberg, S., Savolainen, M.J., Verhoeven, A., Viikari, J., Willemsen, G., Boomsma, D.I., van Duijn, C.M., Eriksson, J., Jula, A., Järvelin, M.R., Kaprio, J., Metspalu, A., Raitakari, O., Salomaa, V., Slagboom, P.E., Waldenberger, M., Ripatti, S., Ala-Korpela, M., 2016. Genome-wide study for circulating metabolites identifies 62 loci and reveals novel systemic effects of LPA. Nat. Commun. 7, 11122.

[19] Liu, Y., Basty, N., Whitcher, B., Bell, J.D., Sorokin, E.P., van Bruggen, N., Thomas, E.L., Cule, M., 2021. Genetic architecture of 11 organ traits derived from abdominal mri using deep learning. Elife 10, e65554.

[20] Liu, Y., Lai, H., Zhang, R., Xia, L., Liu, L., 2023. Causal relationship between gastro-esophageal reflux disease and risk of lung cancer: insights from multivariable Mendelian randomization and mediation analysis. International Journal of Epidemiology, dyad090URL: https://doi.org/10.1093/ije/dyad090, doi:10.1093/ije/dyad090.

[21] Loos, R.J.F., Yeo, G.S.H., 2022. The genetics of obesity: from discovery to biology. Nat. Rev. Genet. 23, 120–133.

[22] McPherson, R., Tybjaerg-Hansen, A., 2016. Genetics of coronary artery disease. Circulation Research 118, 564–578.

[23] Nurk, S., Koren, S., Rhie, A., Rautiainen, M., Bzikadze, A.V., Mikheenko, A., Vollger, M.R., Altemose, N., Uralsky, L., Gershman, A., Aganezov, S., Hoyt, S.J., Diekhans, M., Logsdon, G.A., Alonge, M., Antonarakis, S.E., Borchers, M., Bouffard, G.G., Brooks, S.Y., Caldas, G.V., Chen, N.C., Cheng, H., Chin, C.S., Chow, W., de Lima, L.G., Dishuck, P.C., Durbin, R., Dvorkina, T., Fiddes, I.T., Formenti, G., Fulton, R.S., Fungtammasan, A., Garrison, E., Grady, P.G.S., Graves-Lindsay, T.A., Hall, I.M., Hansen, N.F., Hartley, G.A., Haukness, M., Howe, K., Hunkapiller, M.W., Jain, C., Jain, M., Jarvis, E.D., Kerpedjiev, P., Kirsche, M., Kolmogorov, M., Korlach, J., Kremitzki, M., Li, H., Maduro, V.V., Marschall, T., McCartney, A.M., McDaniel, J., Miller, D.E., Mullikin, J.C., Myers, E.W., Olson, N.D., Paten, B., Peluso, P., Pevzner, P.A., Porubsky, D., Potapova, T., Rogaev, E.I., Rosenfeld, J.A., Salzberg, S.L., Schneider, V.A., Sedlazeck, F.J., Shafin, K., Shew, C.J., Shumate, A., Sims, Y., Smit, A.F.A., Soto, D.C., Sovic, I., Storer, J.M., Streets, A., Sullivan, B.A., Thibaud-Nissen, F., Torrance, J., Wagner, J., Walenz, B.P., Wenger, A., Wood, J.M.D., Xiao, C., Yan, S.M., Young, A.C., Zarate, S., Surti, U., McCoy, R.C., Dennis, M.Y., Alexandrov, I.A., Gerton, J.L., O’Neill, R.J., Timp, W., Zook, J.M., Schatz, M.C., Eichler, E.E., Miga, K.H., Phillippy, A.M., 2022. The complete sequence of a human genome. Science 376, 44–53. doi:10.1126/science.abj6987.

[24] Pierce, B.L., Ahsan, H., VanderWeele, T.J., 2011. Power and instrument strength requirements for mendelian randomization studies using multiple genetic variants. International journal of epidemiology 40, 740–752.

[25] Schunkert, H., König, I.R., Kathiresan, S., Reilly, M.P., Assimes, T.L., Holm, H., Preuss, M., Stewart, A.F.R., Barbalic, M., Gieger, C., Absher, D., Aherrahrou, Z., Allayee, H., Altshuler, D., Anand, S.S., Andersen, K., Anderson, J.L., Ardissino, D., Ball, S.G., Balmforth, A.J., Barnes, T.A., Becker, D.M., Becker, L.C., Berger, K., Bis, J.C., Boekholdt, S.M., Boerwinkle, E., Braund, P.S., Brown, M.J., Burnett, M.S., Buysschaert, I., Cardiogenics Carlquist, J.F., Chen, L., Cichon, S., Codd, V., Davies, R.W., Dedoussis, G., Dehghan, A., Demissie, S., Devaney, J.M., Diemert, P., Do, R., Doering, A., Eifert, S., Mokhtari, N.E.E., Ellis, S.G., Elosua, R., Engert, J.C., Epstein, S.E., de Faire, U., Fischer, M., Folsom, A.R., Freyer, J., Gigante, B., Girelli, D., Gretarsdottir, S., Gudnason, V., Gulcher, J.R., Halperin, E., Hammond, N., Hazen, S.L., Hofman, A., Horne, B.D., Illig, T., Iribarren, C., Jones, G.T., Jukema, J.W., Kaiser, M.A., Kaplan, L.M., Kastelein, J.J.P., Khaw, K.T., Knowles, J.W., Kolovou, G., Kong, A., Laaksonen, R., Lambrechts, D., Leander, K., Lettre, G., Li, M., Lieb, W., Loley, C., Lotery, A.J., Mannucci, P.M., Maouche, S., Martinelli, N., McKeown, P.P., Meisinger, C., Meitinger, T., Melander, O., Merlini, P.A., Mooser, V., Morgan, T., Mühleisen, T.W., Muhlestein, J.B., Münzel, T., Musunuru, K., Nahrstaedt, J., Nelson, C.P., Nöthen, M.M., Olivieri, O., Patel, R.S., Patterson, C.C., Peters, A., Peyvandi, F., Qu, L., Quyyumi, A.A., Rader, D.J., Rallidis, L.S., Rice, C., Rosendaal, F.R., Rubin, D., Salomaa, V., Sampietro, M.L., Sandhu, M.S., Schadt, E., Schäfer, A., Schillert, A., Schreiber, S., Schrezenmeir, J., Schwartz, S.M., Siscovick, D.S., Sivananthan, M., Sivapalaratnam, S., Smith, A., Smith, T.B., Snoep, J.D., Soranzo, N., Spertus, J.A., Stark, K., Stirrups, K., Stoll, M., Tang, W.H.W., Tennstedt, S., Thorgeirsson, G., Thorleifsson, G., Tomaszewski, M., Uitterlinden, A.G., van Rij, A.M., Voight, B.F., Wareham, N.J., Wells, G.A., Wichmann, H.E., Wild, P.S., Willenborg, C., Witteman, J.C.M., Wright, B.J., Ye, S., Zeller, T., Ziegler, A., Cambien, F., Goodall, A.H., Cupples, L.A., Quertermous, T., März, W., Hengstenberg, C., Blankenberg, S., Ouwehand, W.H., Hall, A.S., Deloukas, P., Thompson, J.R., Stefansson, K., Roberts, R., Thorsteinsdottir, U., O’Donnell, C.J., McPherson, R., Erdmann, J., CARDIoGRAM Consortium, Samani, N.J., 2011. Large-scale association analysis identifies 13 new susceptibility loci for coronary artery disease. Nat. Genet. 43, 333–338.

[26] Verbanck, M., Chen, C.Y., Neale, B., Do, R., 2018. Detection of widespread horizontal pleiotropy in causal relationships inferred from mendelian randomization between complex traits and diseases. Nature Genetics 50, 693–698. URL: https://doi.org/10.1038/s41588-018-0099-7, doi:10.1038/s41588-018-0099-7.

[27] Weiss, L.A., Lin Pan, M.A., Ober C., 2006. The sex-specific genetic architecture of quantitative traits in humans. Nature genetics 38, 218—-222.

[28] Weledji, E.P., 2018. Familial hepatocellular carcinoma: ‘a model for studying preventive and therapeutic measures’. Annals of Medicine and Surgery 35, 129–132. URL: https://doi.org/10.1016/j.amsu.2018.09.035, doi:10.1016/j.amsu.2018.09.035.

[29] Wilman, H.R., Parisinos, C.A., Atabaki-Pasdar, N., Kelly, M., Thomas, E.L., Neubauer, S., Jennison, C., Ehrhardt, B., Baum, P., Schoelsch, C., et al., 2019. Genetic studies of abdominal MRI data identify genes regulating hepcidin as major determinants of liver iron concentration. Journal of Hepatology 71, 594–602. URL: https://doi.org/10.1016/j.jhep.2019.05.032, doi:10.1016/j.jhep.2019.05.032.

[30] Yucesoy, B., Charles, L.E., Baker, B., Burchfiel, C.M., 2015. Occupational and genetic risk factors for osteoarthritis: a review. Work 50, 261–273.

